# Shared loci but distinct variants underlie genetic architecture of allergic diseases

**DOI:** 10.1101/2025.06.06.25329154

**Authors:** Isabella M. Salamone, Xiaoyuan Zhong, Noboru J. Sakabe, Robert Mitchell, William Wentworth-Sheilds, Li Zhang, Emma E. Thompson, Donna C. Decker, Clayton H. Rische, Xuanyao Liu, Nathan Schoettler, Carole Ober, Xin He, Marcelo A. Nóbrega

## Abstract

Asthma, allergic rhinitis, and atopic dermatitis are common, complex traits that are frequently co-morbid and have strong genetic correlation. However, the extent to which genome-wide genetic correlation between traits reflects shared causal variants or risk genes remains unclear. To address this question, we used functional fine-mapping. We generated genomic annotations from primary cells treated with immunomodulatory stimuli, then used these data to identify likely causal variants mediating genetic risk for allergic diseases including adult-onset asthma, childhood-onset asthma, allergic rhinitis, and atopic dermatitis. After identifying likely causal variants, we combined our functional annotations with expression quantitative trait loci and activity-by-contact modeling to predict effector genes. We confirmed a high degree of genetic correlation between GWAS loci for allergic diseases, but on the local level very few of the hundreds of likely causal variants identified by functional fine-mapping were shared between diseases. Instead, we found that each allergic disease was associated with a set of mostly unique variants. Nonetheless, nearly 40% of effector genes predicted to be the regulatory targets of these variants were shared between more than one allergic disease. When we tested candidate regulatory elements containing likely causal variants, we found that regulatory elements demonstrated variable allele-specific enhancer activity depending on the cell type in which they were tested. Overall, our findings suggest a highly pleiotropic gene regulatory network underlying allergic diseases, wherein disease-specific risk variants affect different regulatory elements that converge on the same set of target genes.

## INTRODUCTION

Asthma and other allergic diseases are common, chronic diseases with high global prevalence. In the United States, asthma affects over 24 million people; nearly a third of adults and a quarter of all children have seasonal allergies or eczema (*1–4*). While allergic diseases share chronic inflammation as an underlying pathology, there are key features that distinguish them. Atopic dermatitis (AD), or eczema, presents primarily in early childhood as redness, blistering, and rashes of the skin (*5*). Asthma, which can be childhood-onset (COA) or adult-onset (AOA), is a lung disease characterized by wheezing and bronchoconstriction of the airways (*6*). Allergic rhinitis (AR), or hay fever, can develop in late childhood but is most frequent in adults; inflammation of the nasal mucosal membranes occurs in response to either outdoor (pollen) or indoor (dust, pet dander) allergens (*7*). Despite their different clinical presentations and etiologies, allergic diseases often co-occur; approximately 80% of patients with asthma have allergic rhinitis, up to 38% of patients with allergic rhinitis have asthma, and about 30% of children with atopic dermatitis develop asthma, while about 66% develop allergic rhinitis (*8*). While the risk of developing allergic diseases is known to be influenced by both genetic and environmental factors (*9–11*), how these factors might lead to the development of one versus multiple allergic diseases is poorly understood. Data from genome-wide association studies (GWAS) have pinpointed genomic regions with genetic correlations between allergic diseases, suggesting the existence of shared risk loci, as well as unique signals that might contribute to the distinct pathology of an individual allergic disease (*12*). However, because there is substantial correlation between variants in close proximity, i.e. linkage disequilibrium (LD), it is not clear which variants are causal at each locus. It remains an open question whether causal risk variants are shared between allergic diseases and the extent to which genetic correlation reflects shared causal variants.

The aim of this study was to address these questions by interrogating the genetic architecture of allergic diseases beyond the level of genome-wide association. Fine-mapping uses statistical modeling to identify the variants at a GWAS locus that are likely to be causal. Previous allergic disease fine-mapping studies have identified a small number of pleiotropic variants shared between asthma, AR, and AD (*13*), allergic and autoimmune diseases (*14*), and a wide spectrum of allergic diseases (*15*). We recently applied a functional fine-mapping approach to improve detection of causal variants mediating AOA and COA genetic risk (*16*). Functional fine-mapping leverages genomic annotations such as chromatin accessibility to identify causal variants, with the rationale that a variant mapping within an open chromatin region (OCR) is more likely to be functionally consequential and accordingly causal than a variant that does not. We identified a small number (16%) of credible sets shared between AOA and COA despite their high genetic correlation. To directly investigate the extent to which allergic disease risk is mediated by pleiotropic versus unique risk variants, we used functional fine-mapping to identify the likely causal variant(s) at each GWAS locus for AOA, COA, AR, and AD. We identified strong genetic correlation and substantial overlap in LD blocks containing regions with genome-wide significant association between allergic diseases, but out of 2,096 fine-mapped variants, only 152 were shared between more than one allergic disease. Intriguingly, both pleiotropic and unique variants converged on a similar set of risk genes, suggesting that asthma and allergic disease genetic risk is mediated by disease-specific variation that regulates a core set of conserved cellular and molecular processes.

## RESULTS

### Genome-wide functional annotations in primary human cells

We first generated comprehensive genomic annotations of primary cell types involved in the pathogenesis of allergic diseases (Figure 1). In addition to data collected from primary bronchial epithelial cells (BECs) that we have previously described (*17*), we generated comprehensive transcriptomic (RNA-seq), chromatin accessibility (ATAC-seq), and long-range chromatin interaction (PCHi-C) datasets from primary airway smooth muscle cells (ASMCs) and alveolar (AMs) and interstitial (IMs) macrophages (Supplementary Table 1). Development and exacerbation of asthma and other allergic diseases can result from exposure to environmental stimuli such as viruses and microbial infections, as well as inflammatory cytokines produced by the allergic milieu (*18–21*). To detect both constitutively open and dynamically regulated genes and OCRs, we treated ASMCs with IL-13 or IL-17 and both AMs and IMs with lipopolysaccharide (LPS). We identified 142,658 and 166,648 OCRs in AMs and IMs respectively and 174,177 OCRs in ASMCs (Figure 2A-B). We had previously identified 57,147 OCRs in BECs exposed to rhinovirus (RV) (*17*). We also identified 240,903 chromatin-promoter interactions in AMs and IMs and 2,552,190 interactions in ASMCs, in addition to the 601,845 interactions we previously identified in BECs (*17*). Treatment with LPS resulted in 2,270 and 2,296 differentially expressed genes (DEGs) in AMs and IMs, respectively (Supplementary Figure 1). DEGs in both macrophage subsets were enriched for inflammatory cytokine and TNF signaling pathways (Supplementary Figure 1). ASMCs treated with IL-13 had 2,181 DEGs that were enriched for cell cycle and DNA replication pathways, consistent with the known function of IL-13 as a proliferation inhibitor (*22*). Unlike the broad differences in gene expression following IL-13 stimulation, we observed a limited number of DEGs (n = 137) after IL-17 treatment in ASMCs, similar to results reported by others (*23*). To define possible upstream regulators mediating the dynamic responses we observed, we used HOMER to identify transcription factor binding motifs that were enriched in chromatin regions that opened or closed in each cell type after treatment (Figure 2C). The most significant motifs enriched in dynamic OCRs in all cell types included AP-1 members Fos and Jun, reflecting this transcription factor’s key role in mediating the cellular response to cytokines and bacterial and viral infections (*24*). The genes encoding these transcription factors were differentially expressed in AMs and IMs after LPS, ASMCs after IL-13, and BECs after RV (Figure 2D). OCRs opening in ASMCs after IL-13 were uniquely enriched for motifs of transcription factors that regulate the cell cycle, including TWIST2 and TCF4, both of which had significantly increased expression after cytokine treatment. Overall, we generated high-quality transcriptomes and chromatin regulatory maps in multiple primary cell types.

**Fig 1.**
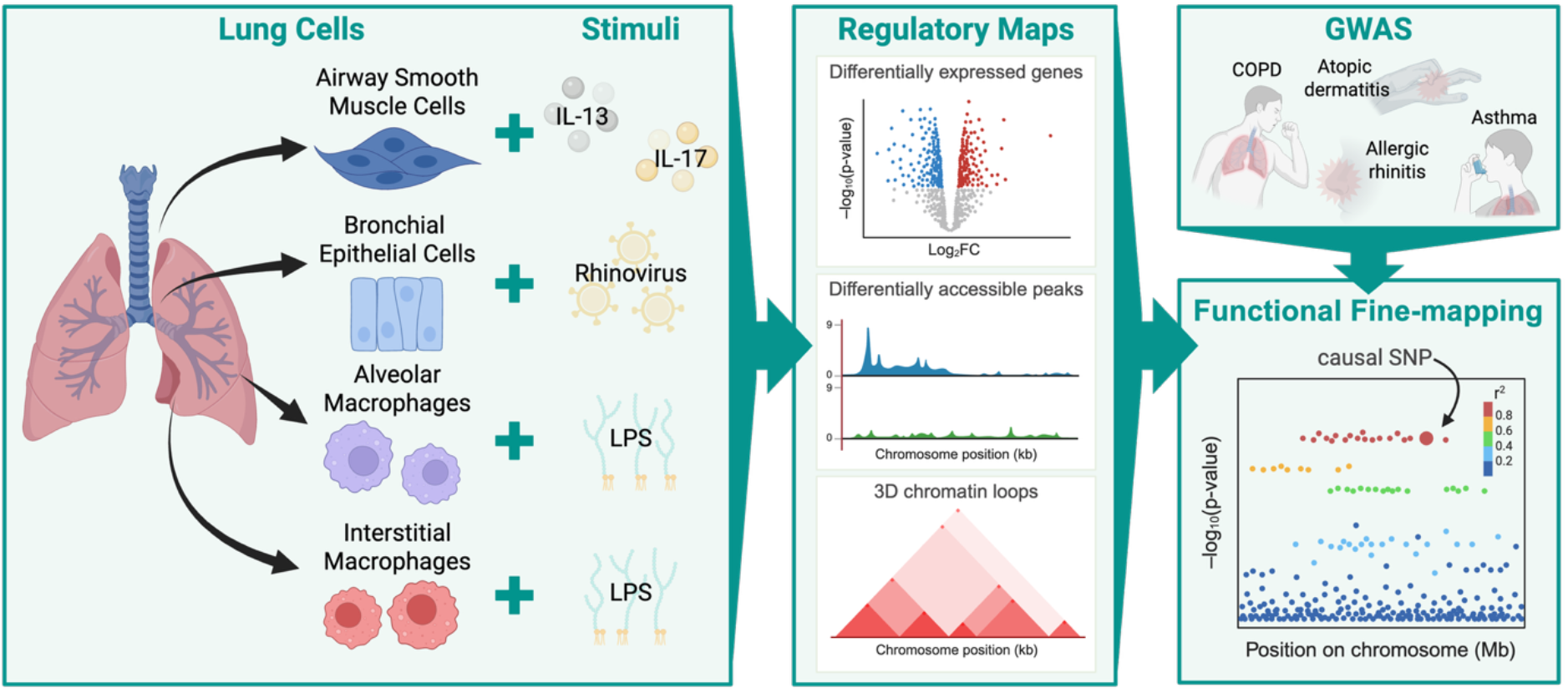
Generation of genomic annotations. Primary airway smooth muscle cells, bronchial epithelial cells, alveolar macrophages, and interstitial macrophages isolated from donor lungs were stimulated in culture. Regulatory maps including RNA-seq, ATAC-seq, and promoter-capture Hi-C were generated, then used to perform functional fine-mapping on genome-wide association studies for COPD, atopic dermatitis, allergic rhinitis, and asthma.

**Fig 2.**
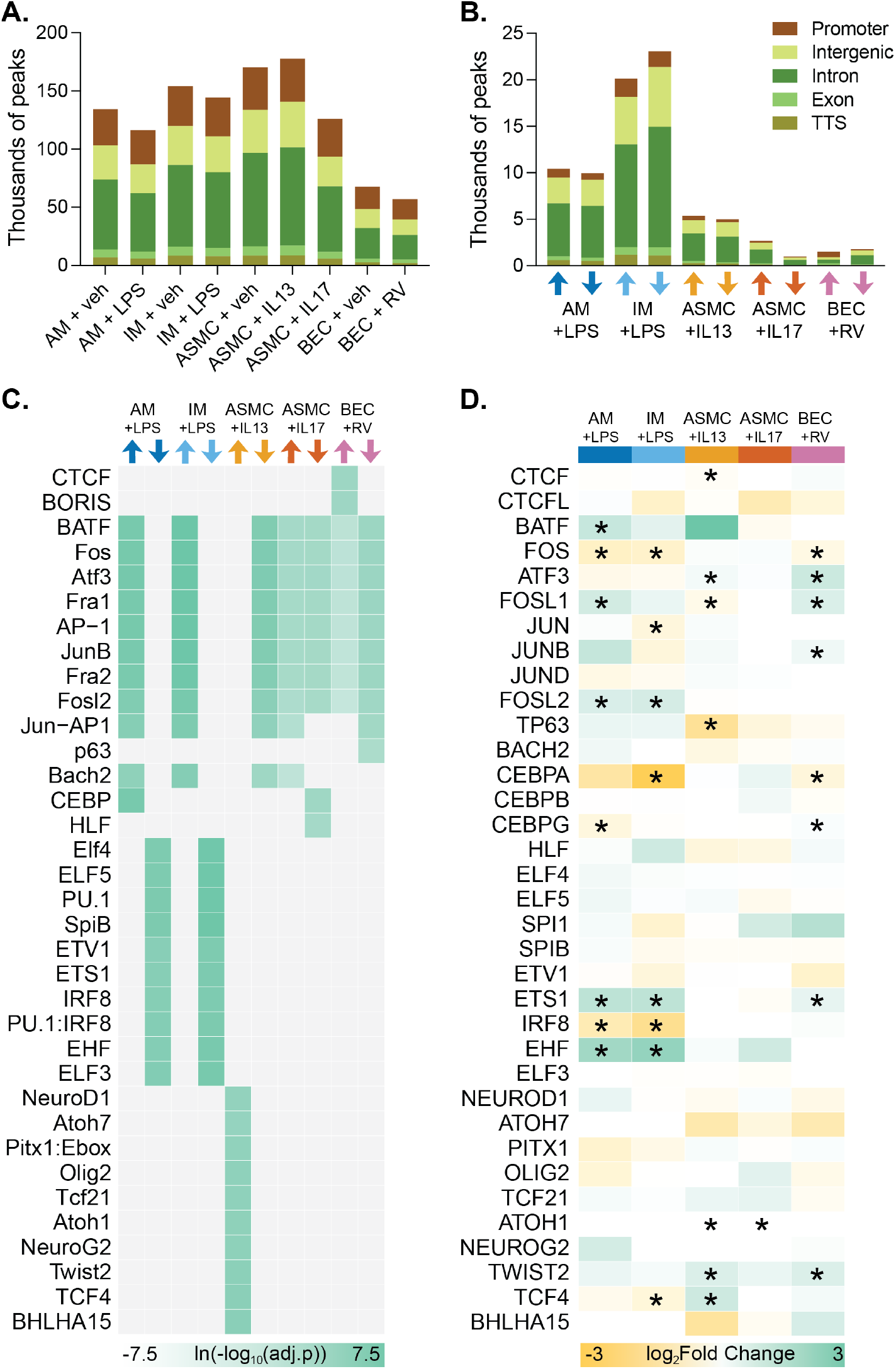
Stimulation of primary cells leads to changes in chromatin accessibility. **(A)** Number of ATAC-seq peaks called in each cell type treated with either vehicle or stimulation. Bar colors represent the genomic feature to which each peak was assigned. **(B)** Number of differentially accessible ATAC-seq peaks that opened (up arrow) or closed (down arrow) after stimulation. **(C)** The top ten enriched transcription factor motifs in each cell type in peaks that opened (up arrow) or closed (down arrow) after stimulation. **(D)** Differential gene expression of transcription factors whose motif was identified in (C), displayed as log2 fold change to vehicle. Asterisks indicate adjusted p-value <0.05.

### Functional fine-mapping of asthma and allergic disease GWAS loci

To investigate the shared genetic architecture of allergic diseases with related respiratory and immune traits, we first used bivariate LD score regression (*25*) to estimate genetic correlations across 12 phenotypes. These included AOA, COA, AR, AD, lung function (first second of force expiration to forced vital capacity (FEV1/FVC)), two lung diseases (chronic obstructive pulmonary disease (COPD), idiopathic pulmonary fibrosis (IPF)), two autoimmune diseases with strong epithelial cell compartments (Crohn’s disease (CD), ulcerative colitis (UC)), and blood biomarkers of inflammation (eosinophil, monocyte, and neutrophil counts) (Figure 3A, Supplementary Table 6). We observed significant positive genetic correlations amongst AOA, COA, AR, and AD, as has been previously reported (*13–15, 26, 27*). Both AOA and COA were positively correlated with COPD and negatively correlated with FEV1/FVC, consistent with the inverse relationship between lung function and COPD status. Despite the presence of shared inflammatory features, the only significant genetic correlation between inflammatory bowel disease and allergic disease was between Crohn’s disease and AD, perhaps due to the compromised epithelial barrier underlying both diseases (*28*). Among three blood cell count traits, eosinophil count was significantly correlated with all allergic diseases, reflecting the key role of eosinophilia in allergic inflammation (*29*), but much less so with other traits.

**Fig 3.**
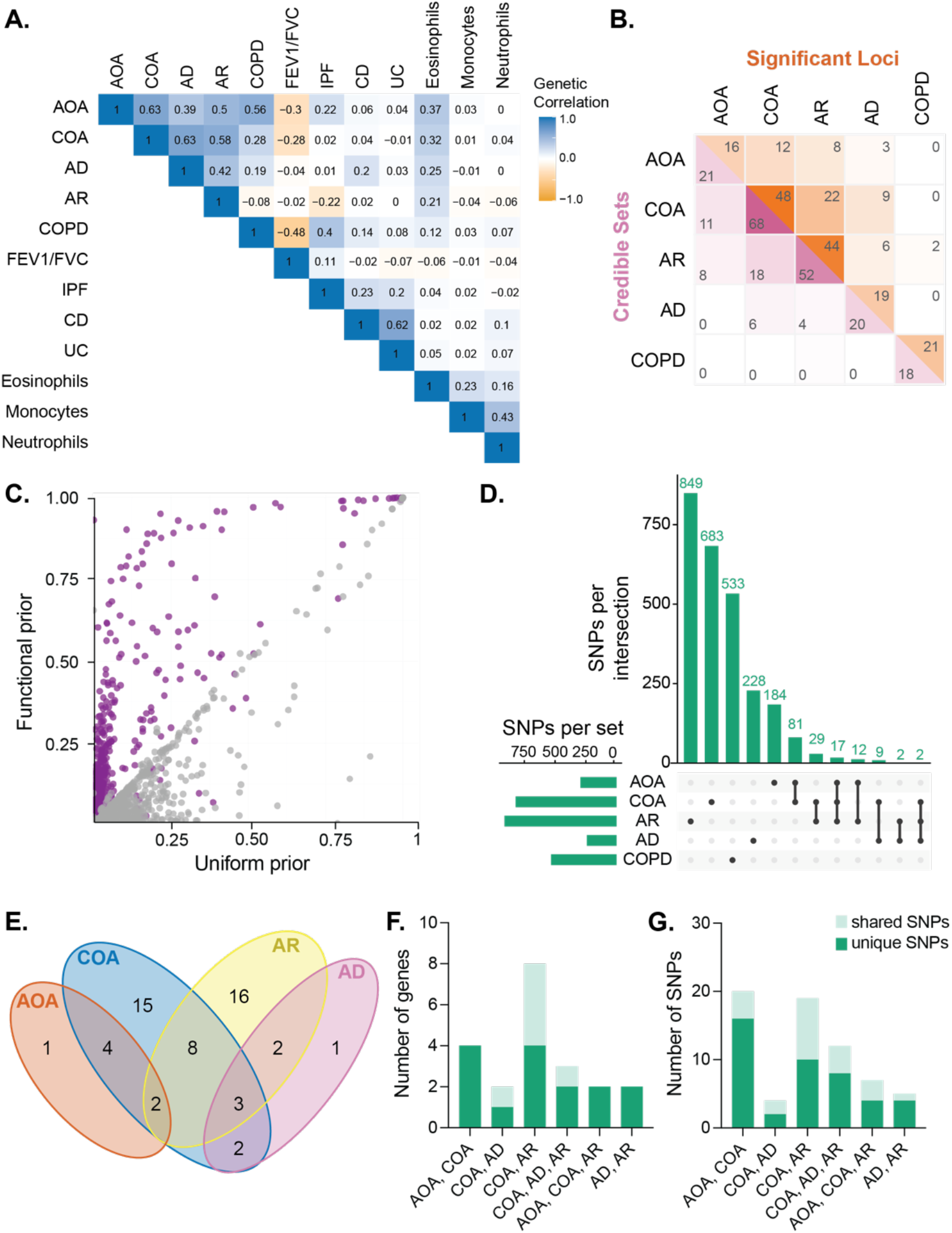
Fine-mapping of respiratory and allergic diseases finds some shared but many unique genetic features. **(A)** Genetic correlation between publicly available GWAS for respiratory, allergic, and immune traits. **(B)** Number of shared significant loci (orange) and credible sets (pink) between respiratory and allergic diseases. **(C)** PIPs for SNPs with (purple) or without (grey) functional annotations in fine-mapping performed under a functional versus uniform prior. **(D)** Number of SNPs were shared between AOA, COA, AR, AD, or COPD. **(E)** Number of genes with a risk score > 0.95. **(F)** Number of high-scoring genes shared between indicated traits (x-axis) that had shared (light green) or unique (dark green) risk variants. **(G)** Number of shared or unique risk variants contributing to high-scoring genes shared between indicated traits (x-axis).

Genetic correlation measures the overall genetic similarity between different traits. To refine our understanding of the shared genetic components at individual loci, we statistically fine-mapped the traits with the strongest genetic correlations: COA, AOA, COPD, AR, and AD. As fine-mapping meta-analyzed GWAS data is prone to false positives (*30*), we first performed GWAS of AD, AR, and COPD using European-ancestry individuals in the UK Biobank (Supplementary Figure 2). We had conducted GWAS of AOA and COA from the same source in our previous study (*16*). As anticipated, we observed overlap in cases with different allergic diseases, the largest being between AOA and AR (n = 4,059, Supplementary Table 2). To perform fine-mapping, we first identified LD blocks containing at least one significant SNP, termed significant loci, for each trait. Consistent with the observed genetic correlation patterns, most significant loci were shared between pairs of allergic diseases (Figure 3B, upper half). In contrast, COPD shared only two significant loci with AR and none with COA, AOA, or AD. Next, we fine-mapped significant loci for each trait using SuSiE-RSS (*31, 32*). To adjust the prior probability of each individual variant being causal, we integrated OCRs from our regulatory maps as well as publicly available single cell ATAC-seq datasets of blood and lung cells (*33, 34*). Using this functional fine-mapping approach, we identified credible sets for each trait, ranging from 18 (COPD) to 68 (COA) (Figure 3B, lower half). As a percent of the total number, there were fewer shared credible sets between traits than shared significant loci; that is, traits that shared significant GWAS loci had different fine-mapped credible sets, and thus likely causal variants, at some loci. Compared to fine-mapping with a uniform prior, SNPs located in OCRs tend to have higher posterior inclusion probabilities (PIPs) under functional fine-mapping (Figure 3C), suggesting our approach effectively prioritizes variants within candidate regulatory elements.

We next investigated the sharing of likely causal SNPs between traits. In total, we identified 294, 821, 911, 241, and 533 putative causal variants for AOA, COA, AR, AD, and COPD, respectively (Figure 3D). Surprisingly, the vast majority of variants were specific to individual traits. The highest overlap was between AOA and COA (n = 81 SNPs) and no variants were shared across more than three traits. Notably, COPD did not share any putative causal variants with any allergic diseases, supporting the notion that global genetic similarity does not necessarily translate to shared genetics at specific loci. Based on this observation, we did not include COPD in the following analyses.

Although fine-mapping identified largely different sets of causal variants for different traits, we hypothesized that distinct variants at shared loci might converge on the same set of causal genes. To investigate effector gene sharing across allergic diseases, we used both coding variants and fine-mapped non-coding variants to nominate putative causal genes for each trait. For non-coding variants, we used genomic annotations including our PCHi-C maps in primary cells, expression quantitative trait loci (eQTL), and activity-by-contact models to link variants to putative target genes. We then calculated a risk score for each gene based on the contribution of these linked variants (*16*). We nominated 54 high-confidence risk genes across allergic diseases; 21 genes were shared between at least two traits. We observed substantial overlap in high-scoring genes between AR and COA (n = 13 genes) and modest overlap between the other diseases (Figure 3E, Supplementary Table 4). Intriguingly, although nearly 40% of high-scoring genes were shared between two or more traits, the majority (71.4%) of these genes were supported by at least one risk variant unique to one trait (Figure 3F, 3G). The credible sets of these unique SNPs were largely distinct (Supplementary Table 5). Overall, our findings demonstrate that different variants contribute to disease risk in allergic diseases, suggesting that, at individual loci, different disease-specific regulatory elements converge on the same genes.

### Functional validation of enhancers with tissue-specific activity

To investigate the role cell type might play in the pleiotropic gene regulation that we observed in our fine-mapping analyses, we chose four variants to characterize more deeply. To investigate the potential enhancer function of these variants, we performed luciferase reporter assays in hTERT-immortalized airway smooth muscle (hTERT-ASM) cells, an immortalized monocyte cell line differentiated to macrophages (THP1s), and immortalized bronchial epithelial cells (16HBEs). We defined the putative regulatory element as the OCR containing the variant of interest. To demonstrate enhancer activity, a sequence had to have significantly increased luciferase activity compared to a neutral sequence control (Supplementary Figure 4) and a fold change to the neutral sequence greater than 1.4 (log2 fold change > 0.5) (*35–37*).

Two variants were identified as likely causal SNPs in COA only. The first variant, rs12130219, is located on chromosome 1 overlapping the long non-coding RNA *FLG-AS1* and is an eQTL for *FLG* in lung and immune cells (*38, 39*). We observed luciferase enhancer activity that was both allele- and cell type-specific to the COA protective allele in 16HBEs (Figure 4A). The second variant, rs62279869, is located on chromosome 3 near *LPP* (Figure 4B). rs62279869 overlapped an OCR in all cell types and was linked by PCHi-C loops in ASMCs and BECs to *BCL6*. The locus containing this variant demonstrated enhancer activity in both THP1s and 16HBEs, but not hTERT-ASMs.

**Fig 4.**
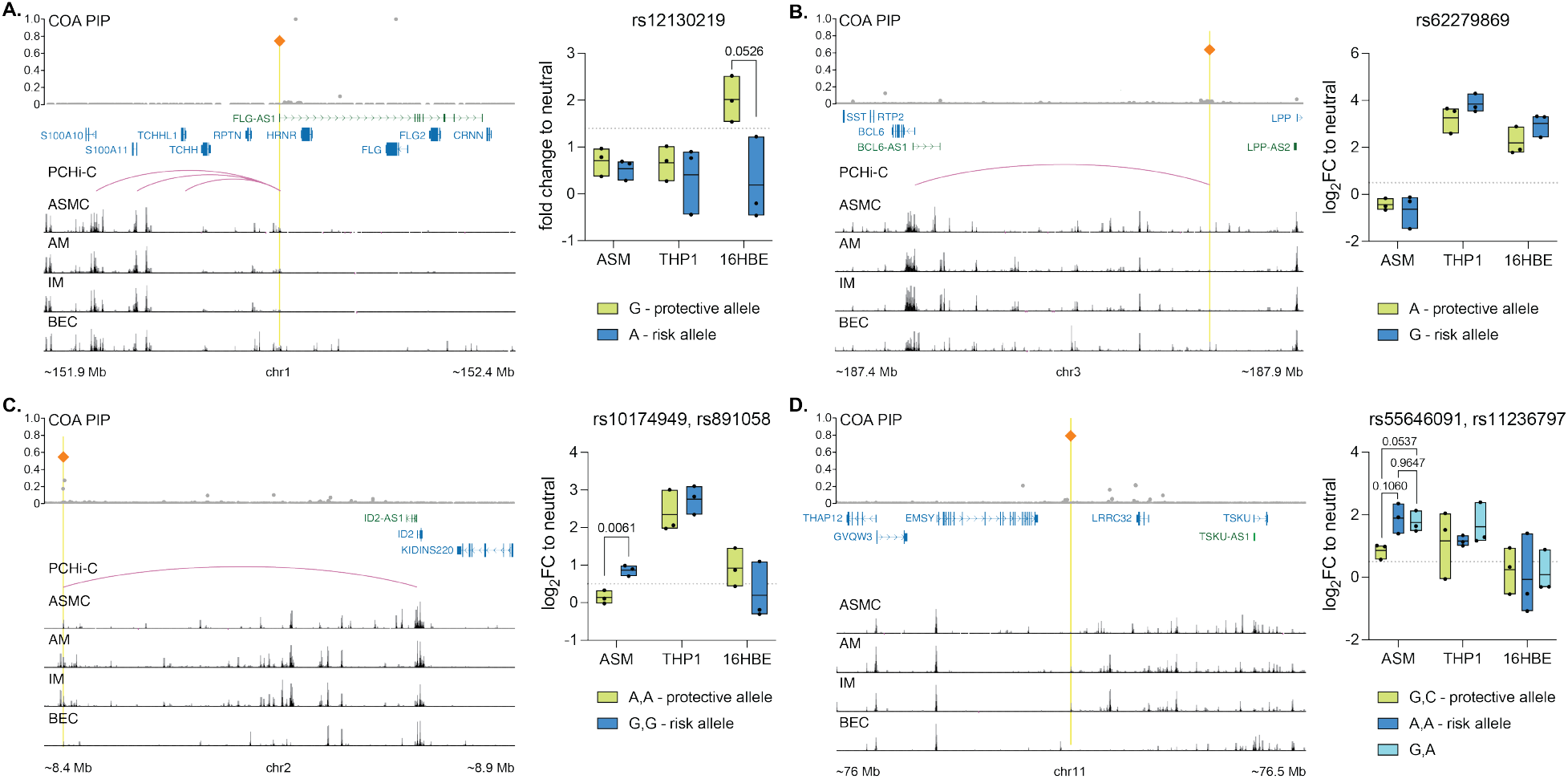
Fine-mapping identifies cell type-specific enhancers. Loci of interest chosen for further characterization: PIP from COA fine-mapping, indicating the likely causal SNP (orange diamond) as the SNP with the highest PIP that overlapped an open chromatin region; coding genes (blue) and long non-coding RNAs (green) at the locus; PCHi-C loops from the causal SNP to promoters at the locus; ATAC-seq peak tracks in primary cells. Yellow line indicates the position of the open chromatin region containing the causal SNP that was assayed for luciferase activity. Open chromatin regions containing putative causal SNPs were assayed for enhancer activity in three cell lines. **(A)** Chr1: rs12130219. **(B)** Chr 3: rs62279869. **(C)** Chr2: rs10174949, rs891058. **(D)** Chr11: rs55646091, rs11236797. A-C, Welch t-test. D, One-way ANOVA.

We also investigated two loci that were identified as likely causal SNPs in more than one allergic disease. One locus, on chromosome 2 near *ID2*, contained two SNPs identified as likely causal variants in both COA and AR (Figure 4C). The two variants, rs10174949 and rs891058, are 299 nucleotides apart and in strong LD in European ancestry populations. These variants overlapped an OCR in AMs, IMs, and ASMCs and were linked by PCHi-C to *ID2* in ASMCs and BECs. The locus containing these SNPs was an enhancer in THP1s and hTERT-ASMs, demonstrating allele-specific regulatory activity in the latter (Figure 4C). A second locus, on chromosome 11 near *LRRC32*, contained two SNPs identified as likely causal variants in COA, AD, and AR (Figure 4D). These variants, rs55646091 and rs11236797, are present in three common haplotypes in European ancestry populations. These variants overlapped an OCR in macrophages. This locus demonstrated enhancer activity in both hTERT-ASMs and THP1s. The two haplotypes containing the risk allele of rs11236797 had slightly higher enhancer activity, suggesting allele-specific activity.

Using fine-mapping and genomic annotations, we identified and characterized a set of candidate enhancers that demonstrate a range of allele- and cell type-specific regulatory activity. These enhancers, harboring candidate causal allergic disease-associated variants, serve as an entry point for the design of variant to function studies in the future that will dissect the cellular and molecular mechanisms underlying the genetic associations in each allergic disease locus.

## DISCUSSION

We generated comprehensive genomic annotations in primary cells and leveraged these regulatory maps to perform functional fine-mapping of loci associated with asthma and other allergic diseases. By intersecting causal variants with OCRs, we were able to identify candidate regulatory elements mediating the heritability of allergic disease. Although we observed substantial overlap of significant loci between asthma and other allergic diseases, the fraction of shared credible sets was surprisingly small, and the fraction of shared likely causal variants smaller still. For example, 45.8% of COA significant loci, 26.4% of credible sets, and 5.8% of likely causal variants were shared with AR (Figure 3). These data suggest that, while these shared genomic regions drive underlying disease risk, the causal variants mediating the risk of each disease are different. While different variants were associated with disease risk in different diseases, these variants often converged on a shared set of genes: 40.6% of highly scored COA risk genes were also highly scored risk genes for AR. Overall, our findings suggest that while allergic diseases share common risk genes and pathways, each trait has a distinct pattern of non-coding genetic regulation that drives disease pathogenesis. This may reflect the organization of the human genome, which contains about 20,000 protein-coding genes and up to a million enhancers. Most individual genes are regulated by a large number of enhancers (*40*), resulting in different variants mediating the activity of separate enhancers on the same gene. This diverse network of regulatory elements allows for context-specific transcriptional regulation in specific tissues, at specific developmental time points, or in response to environmental stimuli (*41*). Under this framework, a distinct set of variants, each associated with different allergic diseases, converge on a small number of common, shared genes, giving rise to the pleiotropism suggested by GWAS (*42, 43*).

Our results suggest that discordance between global and local genetic correlation is likely a feature of the genetic architecture of these traits, and perhaps other complex traits with shared etiologies. Although there is substantial co-morbidity between allergic diseases, there is a significant contribution of environmental factors to the development of each individual trait. Additionally, allergic diseases affect different organs and cell types—the lungs in asthma, skin in AD, and nasal mucosa in AR—which suggests that cell type-specific genetic regulation plays a central role in mediating independent disease risk. The four regulatory elements we experimentally validated with luciferase assays demonstrated enhancer activity in at least one of the tested cell types, but never all three, suggesting that disease-specific variants modulate trait risk through variation of tissue-specific enhancers. We also observed distinct patterns of cell lineage heritability enrichment for each trait (Supplementary Figure 3) and found that only lymphoid enrichment was shared between AOA, COA, AD, and AR, reflecting the important role that B and T cells play in allergic disease (*44, 45*). We speculate that genes with conserved function across traits might be differentially regulated in the context of different disease milieus. For example, we observed that risk gene scoring linked different risk variants to *IL2RA* in AR, AD, and COA. The IL-2 receptor mediates T cell activation as well as type 2 inflammation (*46*). IL-2 dysfunction is a well-described feature of asthma, AR, and AD and IL-2-based therapies have been suggested as a treatment for all three conditions (*47, 48*). However, IL-2 regulation differs between diseases; after allergen challenge, individuals with allergic asthma have significantly different *IL2RA* expression in some lung cell populations compared to allergic individuals without asthma (*49*). It is likely that there are multiple enhancers that regulate *IL2RA* expression, and genetic variation in different regulatory elements mediate disease risk between asthma, AR, and AD. Further investigation beyond the scope of this work is necessary to explore this hypothesis.

Our findings complement and provide nuance to recent work that has used meta-analysis approaches to identify pleiotropic loci, genes, or variants shared between asthma and other allergic diseases (*13–15, 27*). These studies identified shared associations at the gene and locus level, which we also observed as relatively high between allergic diseases. However, there are differences between our approach and previous fine-mapping efforts. We utilized a fine-mapping method that does not assume a single causal variant per locus. Additionally, while fine-mapping of meta-analyzed GWAS data can increase power to detect shared genetic associations, we chose not to work with meta-analyzed data as, without post-processing, it can be prone to nomination of false positive causal signals (*30*). We instead analyzed each trait individually. Our approach thus enabled us to identify risk variants mediating individual unique risk as well as those acting pleiotropically across allergic diseases.

Our study has several limitations. Despite strong association with asthma, AR, and AD, we did not include the HLA region in our fine-mapping studies due to the challenge presented by its high genetic polymorphism, extensive LD, and overall complexity (*50, 51*). We performed individual GWAS for each trait using data from the UKB. Accordingly, our GWAS were conducted in individuals of European ancestry, which may not fully capture the genetic architecture of allergic diseases in non-European populations. A recent cross-population, large-scale fine-mapping study revealed that the genetic architectures of complex traits are highly similar across populations, suggesting that our results may be generalizable to non-European ancestries (*52*). However, we may have missed causal variants common in other ancestries that are rare in Europeans. Due to the substantial rate of allergic disease comorbidity, our individual GWAS each contained a subset of cases that reported more than one allergic disease (Supplementary Table 2). Additionally, differences in sample size across different traits may limit our ability to detect all shared causal variants, potentially leading to an underestimation of genetic similarity at individual loci.

In summary, we have generated comprehensive genomic annotations in primary cells and used these annotations to perform functional fine-mapping for AOA, COA, AR, AD, and COPD. Our work identified pleiotropic regulatory elements and genetic pathways shared between allergic diseases as well as those unique to individual traits. While this work supports the common understanding of allergic diseases as sharing core biological pathways, it also demonstrates a clear distinction between individual allergic disease genetic risk and suggests that pathogenesis of specific allergic diseases is mediated by discrete variants and regulatory elements.

## FIGURES

**Supplementary Fig 1.**
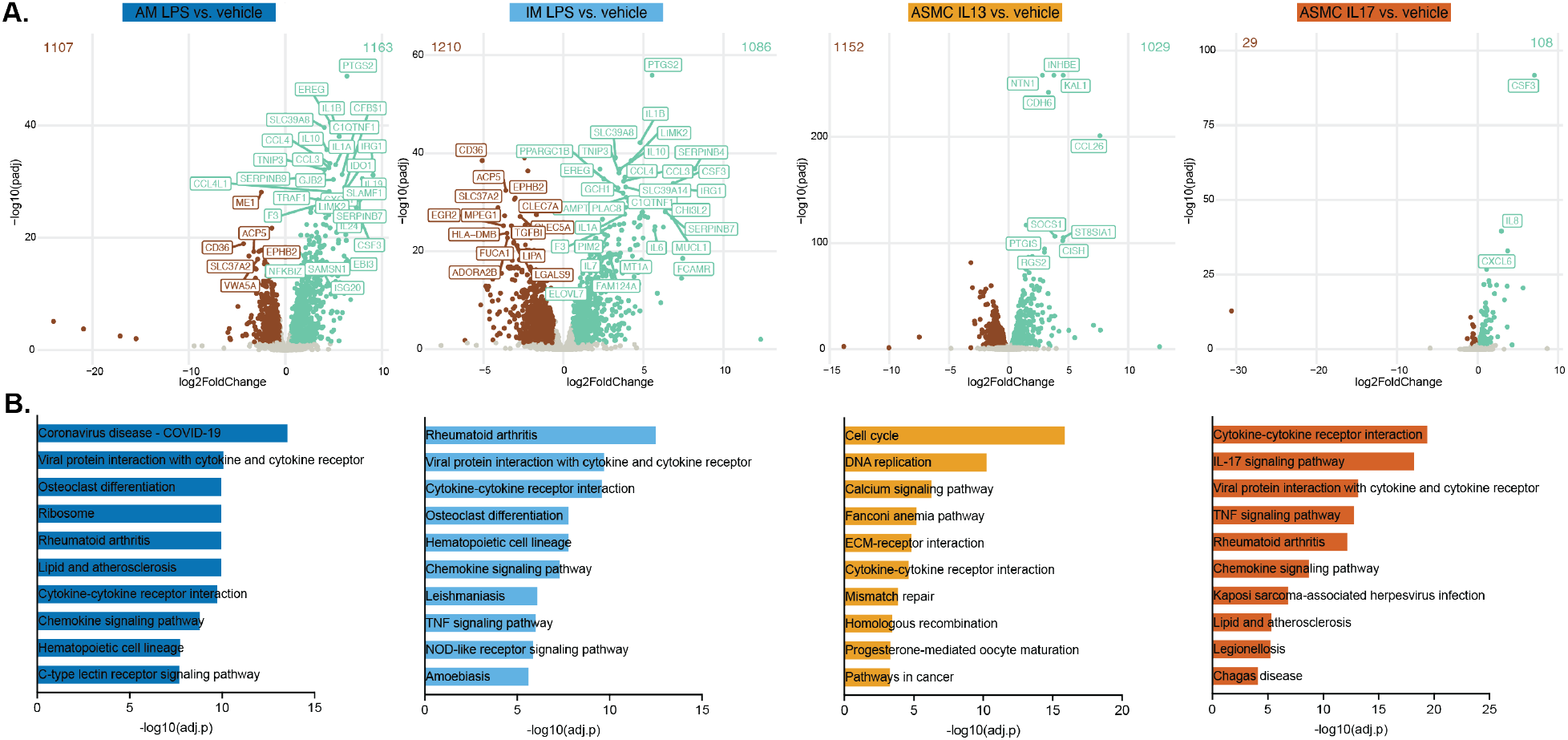
Stimulation of primary cells leads to changes in gene expression. **(A)** Volcano plots showing log2 fold change versus adjusted p-value for differentially expressed genes up-(green) or down-regulated (brown) after stimulation in each cell type. **(B)** Significantly enriched pathways in each cell type after stimulation.

**Supplementary Fig 2.**
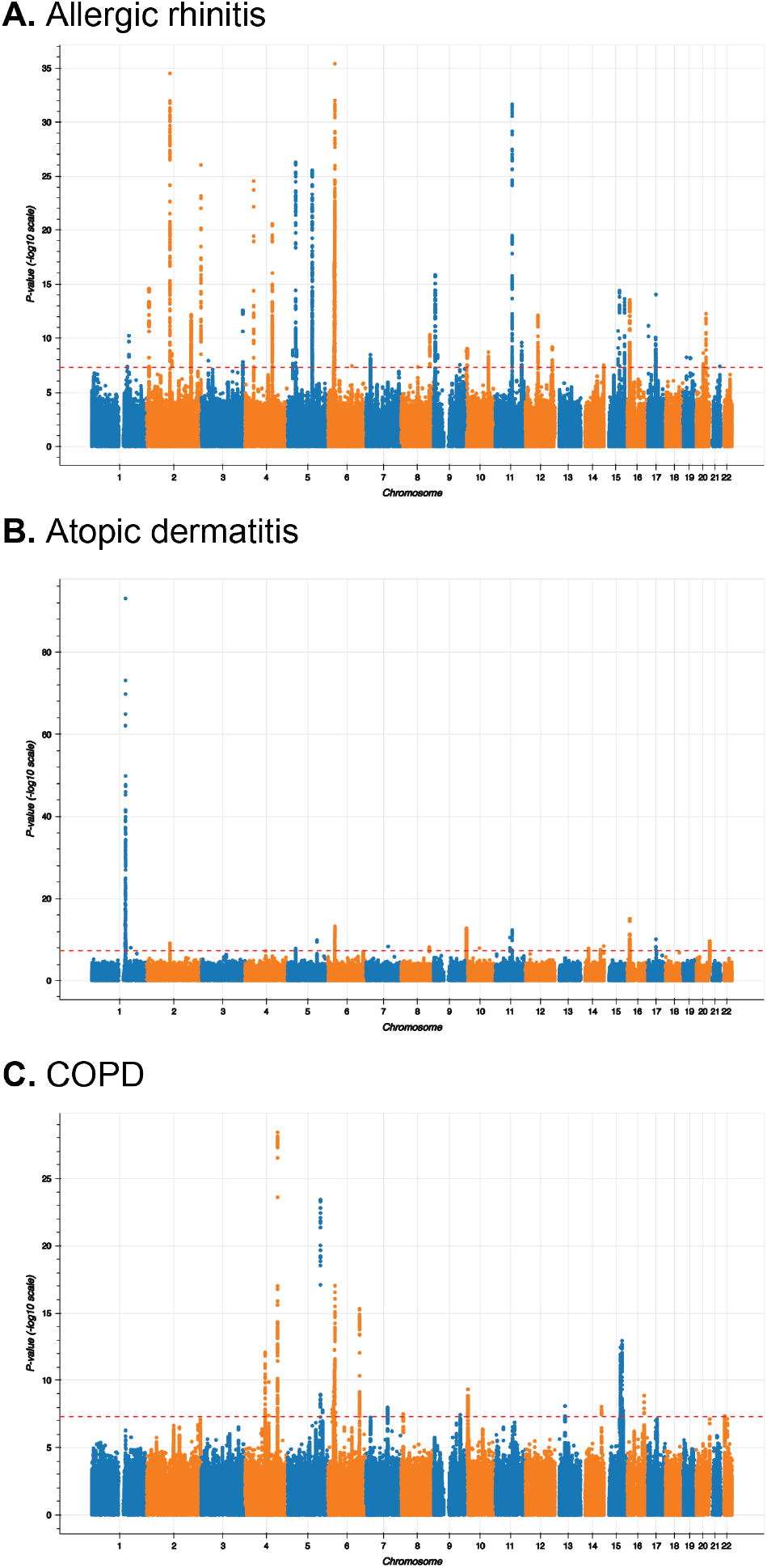
GWAS. Manhattan plots for (A) allergic rhinitis, (B) atopic dermatitis, and (C) COPD.

**Supplementary Fig 3.**
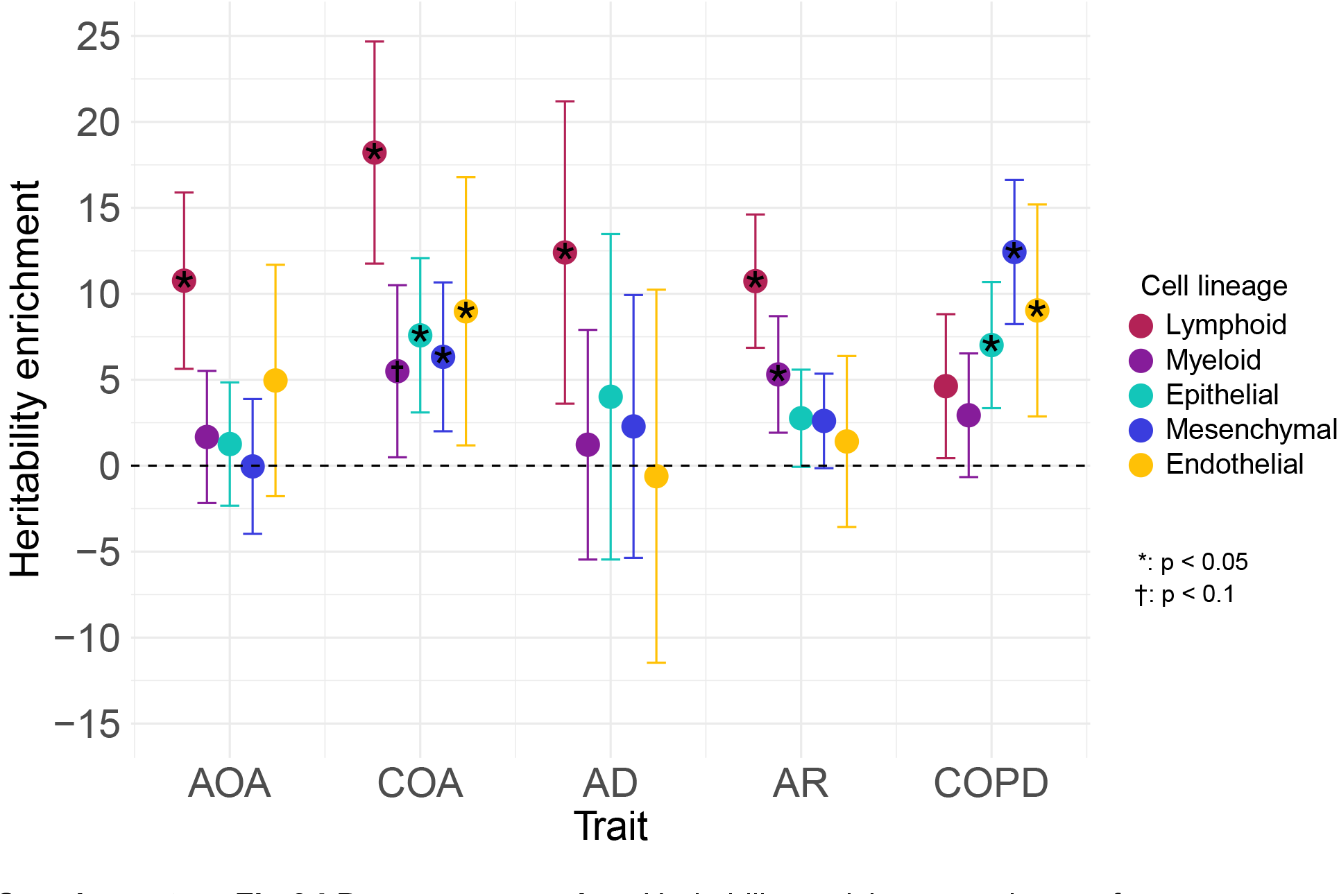
LD score regression. Heritability enrichment estimates for open chromatin regions in allergy-relevant cell types.

**Supplementary Fig 4.**
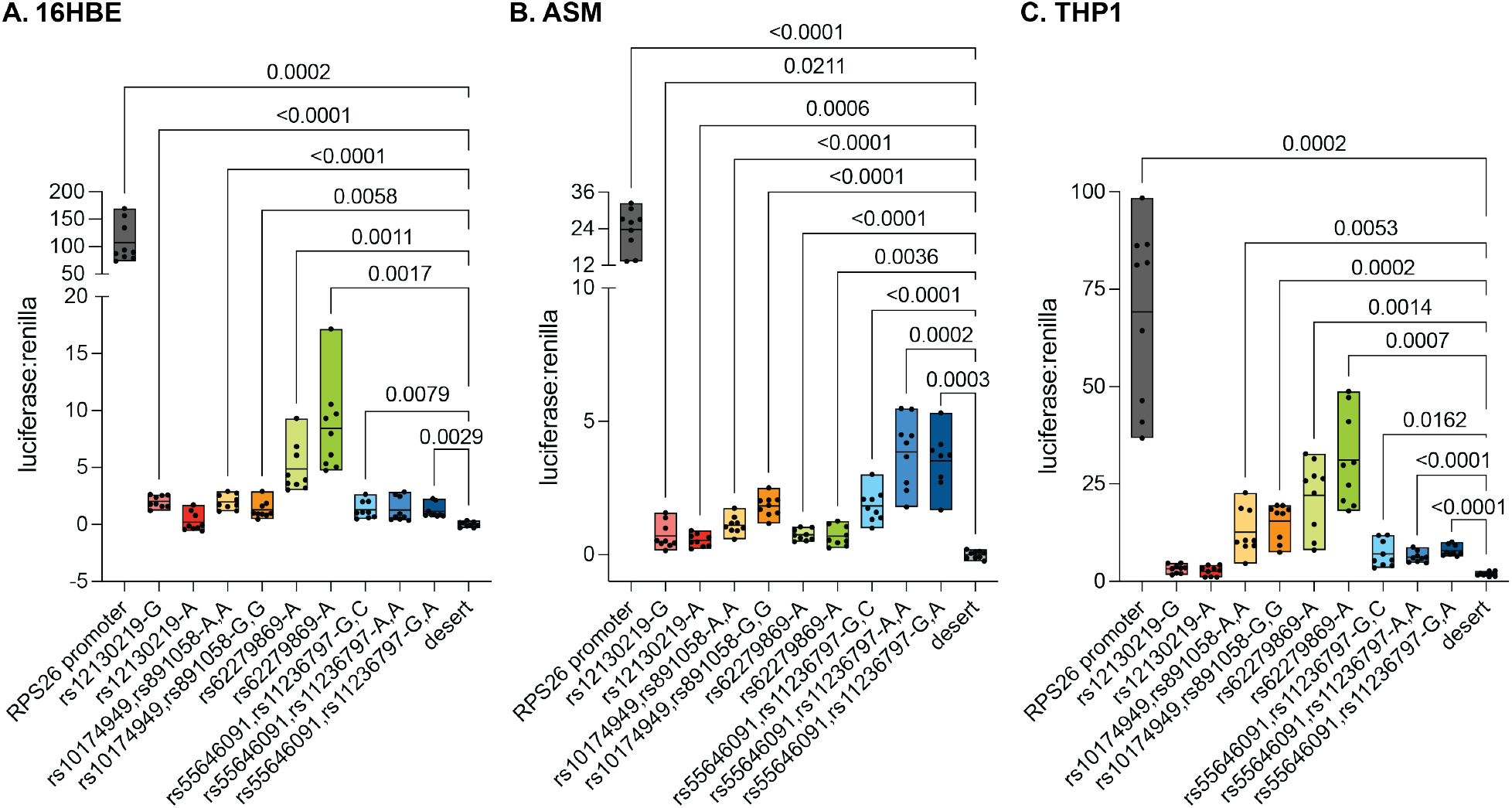
Raw luciferase results identify sequences with enhancer activity. Enhancer activity for each construct reported as luciferase activity normalized to Renilla activity. All constructs, including the positive control (promoter of *RPS26*), were compared to a desert construct negative control. Constructs were transfected into ASMs (A), THP1s (B), or 16HBEs (C) and measured after 24hrs. A-C, Welch ANOVA.

## ACKNOWLEDGEMENTS

This work was supported by the National Institutes of Health under grants U19AI162310 (CO, MAN), R01MH110531, R01HL163523, R01AI175554 (XH), T32HL007605 (IMS), and K08HL153955 (NS).

## METHODS

### Primary cell culture

Primary cells for genomic annotations were isolated from human lungs that were obtained from organ donors whose lungs were not used for transplantation through the Gift of Hope (GOH) Regional Organ Bank of Illinois (ROBI). Donors with more than ten pack-years of tobacco use were excluded from this study. Because samples from this study were from deceased donors, this research was deemed to be non-human subjects research by the Institutional Review Board at the University of Chicago. Donor information is listed in Supplementary Table 1. Cells were processed as previously described (*53*). ASMCs at passage 1 or 2 were cultured in T75 flasks as described in Thompson, Dang, Mitchell-Handley, Rajendran, Ram-Mohan, Solway, Ober and Krishnan (*54*) and treated with IL-13 (10ng/mL), IL-17 (3ng/μL), or vehicle (10% FBS in PBS). Following 24 hours of cytokine exposure, cells for ATAC-seq were trypsinized and counted manually, then separated into aliquots of 50,000 cells for processing. Cells for RNA-seq were trypsinized and RNA was isolated using the QIAgen All-Prep kit. For PCHi-C, 5 million cells were collected from each of two replicates. Macrophages were cultured in RPMI supplemented with 10% FBS and, when noted, separately treated with lipopolysaccharide (LPS; 5μg/ml; eBiosciences, Catalog Number 00-4976). Macrophages were rested for two hours at 37°C prior to the 18-hour LPS treatment. For ATAC-seq and RNA-seq experiments, cells were stained with an antibody panel to distinguish AMs versus IMs: CD16-BV605 (BioLegend 302040), CD45-BUV395 (BD Biosciences, Catalog Number 563792), CD14-PECF594 (BD Biosciences 562335), CD206-PE (BioLegend 321106), CD169-APC (BioLegend 346008) and Apotracker (Biolegend 427402) to ensure high viability. Apotracker-negative cells were sorted on a Symphony S6 cell sorter (BD Biosciences) with gating on the macrophage cells using forward- and side-scatter and the staining panel. IMs were sorted using CD45+CD16+/−CD14+CD206+CD169^lo^/-gates. AMs were sorted with CD45+CD16+/−CD14-CD206+CD169^hi^ gates. For PCHi-C, cells were thawed, processed, and sorted for Apotracker-negative cells. BECs used for ATAC-seq were cultured as described in Helling, Sobreira, Hansen, Sakabe, Luo, Billstrand, Laxman, Nicolae, Nicolae, Bochkov, Gern, Nobrega, White and Ober (*17*).

### Immortalized cell culture

Luciferase assays were performed in human immortalized bronchial epithelial cells (16HBE14o-; Millipore-Sigma SCC150), D9 human telomerase reverse transcriptase immortalized human airway smooth muscle cells (ASMs, generously provided by R. Krishnan and S. Liggett), and human acute monocytic leukemia cells (THP1s, ATCC TIB-202). 16HBEs were cultured as previously described (*16*). ASMs were cultured as previously described (*55, 56*). THP1s were cultured in RPMI with 10% FBS and 0.9μL/mL β-mercaptoethanol. 24 hours before luciferase data was collected, THP1 media was supplemented with 150mM phorbol 12-myristate 13-acetate (Sigma P8139) to induce macrophage differentiation.

### ATAC-seq

ATAC-seq for AMs and IMs was performed according to Corces, Buenrostro, Wu, Greenside, Chan, Koenig, Snyder, Pritchard, Kundaje, Greenleaf, Majeti and Chang (*57*) with minor modifications. Briefly, cell concentration was estimated from freshly sorted cells; approximately 20,000 cells were used for most libraries. Tagmentation was performed in a 50μl reaction containing 25μl 2× TD buffer, 2.5μl TDE1 (Illumina, 20034198), 0.5μl 1% digitonin, and 22μl nuclease-free water. Tagmentation was terminated and DNA collected using Zymo Clean and Concentrator-5 following the manufacturer’s recommendations. Libraries were barcoded with 5 cycles of preamplification, and the PCR reaction was paused for quantification using NEBNext Library Quant Kit for Illumina. Additional cycles were ran as needed to produce at least 10nM of final library. Purified ATAC-seq libraries were sequenced on an Illumina Nova-Seq using paired end 50bp reads with a target depth of 50M reads/sample.

ATAC-seq for ASMCs and BECs was performed as described by Grandi, Modi, Kampman and Corces (*58*). Briefly, nuclei were isolated from 50,000 cells in 50μl ice-cold lysis buffer (10 mM Tris–HCl pH 7.5, 10 mM NaCl, 3 mM MgCl2, 0.1% NP40, 0.1% Tween-20 and 0.01% digitonin) for 3min on ice. Tagmentation was performed on permeabilized nuclei resuspended in 50μl transposition master mix (25μl (2X) TD buffer, 16.5μl PBS, 5μl H2O, 0.5μl 1% digitonin, 0.5μl 10% Tween-20, and 2.5μl Illumina TDE1 enzyme (Tn5 transposase)). Tagmentation was terminated and libraries were prepared and sequenced as described above.

### ATAC-seq peak calling and differential peak analysis

Paired-end ATAC-seq reads were aligned with bowtie2 version 2.3.4.3 (*59*). Reads with mapping quality lower than 10 were discarded. Peaks were called using MACS2 version 2.2.7.1 (*60*) with parameters --llocal 20000 --shift -100 --extsize 200 -q 0.05 for ATAC-seq. MACS 2.2.8 was used for BEC samples. Peaks overlapping coordinates blacklisted by Anshul Kundaje were excluded. Peaks were called for each sample using MACS2 and peak coordinates were converted into 50bp contiguous bins. Bins covered by less than 60% of their extension were excluded. To identify reproducible peaks, we only kept bins that were present in at least N out of D donors in each condition, allowing for condition-specific peaks (ASMC: N = 4, D = 4; alveolar macrophages: N = 4, D = 8, interstitial macrophages: N = 4, D = 11; BEC: N =, D =). We then merged all adjacent bins, expanding them back into longer peaks. We counted the number of reads in all peaks and in all samples and compared the read counts using DESeq2 (ASMC: p-value < 0.1, absolute fold-difference > 1.5; BEC: unadjusted p-value < 0.1 and absolute fold-difference > 1.3, macrophages: adjusted p-value < 0.05 and absolute fold-difference > 1.5).

### RNA-seq

RNA was isolated from snap-frozen cell pellets using Qiagen RNeasy Plus Micro kit with on-column DNase digestion (Qiagen, 74034). AM and IM libraries were generated using the NEB Single Cell/Low Input RNA Library Prep Kit for Illumina (NEB, E6420) following the Protocol for Low Input RNA. ASMC mRNA was isolated using NEBNext Poly(A) mRNA Magnetic Isolation Module (NEB, E7490) and libraries were prepared using NEBNext Ultra II Directional RNA Library Prep Kit for Illumina (NEB, E7760). Sequencing was performed using an Illumina Nova-Seq using paired end 50bp reads with a target depth of 20M reads/sample.

### Differential gene expression

Read counts per gene were obtained with Salmon version 1.5.1 (*61*) on transcripts from human Gencode release 19 (ftp://ftp.ebi.ac.uk/pub/databases/gencode/Gencode_human/release_19/gencode.v19.pc_transcripts.fa.gz and ftp://ftp.ebi.ac.uk/pub/databases/gencode/Gencode_human/release_19/gencode.v19.lncRNA_transcripts.fa.gz). Estimated counts were used in exploratory analysis (transformed with DESeq2’s rlog function) and in DESeq2 version 1.32.0 (*62*) to identify differentially expressed genes (adjusted p-value ≤ 0.05 and absolute fold-change of ≥ 1.2). DESeq2 1.40.0 was used in BEC. An analysis pairing samples by donor (~treatment + donor) was conducted in all datasets (ASMC+IL-13, ASMC+IL-17, AM+ LPS, and AM+LPS).

### Promoter capture Hi-C

In situ high-resolution chromatin conformation capture (Hi-C) was performed according to Lieberman-Aiden et al. 2009 with modifications (see Rao, S. S. P. et al. 2014). Sorted macrophages were mixed 1:1 with 2% formaldehyde (Pierce 28906) in PBS at RT with gentle rotation for 15min. ASMCs were fixed in 1% formaldehyde PBS (pH 7.4) for 10min at RT. Following cell lysis, restriction digestion, proximity ligation, and DNA sheering and size selection, purified Hi-C libraries were with 2.5μ Cot-1 (Invitrogen 15279-001), 2.5μL Salmon Sperm (Invitrogen 15632-001) and 0.5μL each of p5 and p7 blocking primers (IDT 1016184 & 1016186) in hybridization buffer (25μL 20X SSPE (Invitrogen AM9767), 1μL 0.5M EDTA (Invitrogen AM9260), 10μL 50X Denhardt’s Solution (Invitrogen 750018) and 13μL 1% SDS (Invitrogen 15553-035)). Purified libraries were hybridized with 500ng biotinylated RNA oligo bait library for 26h @65°C. Following hybridization, the hybridized probes were washed and resuspended in 21μL water and amplified on-bead to produce the final capture library. Following purification and QC the library was sequenced with Illumina Nova-Seq using paired end 50bp reads and a target depth of 200M reads.

### Promoter capture Hi-C interaction calling

We used HiCUP v0.5.9 (*63*) with bowtie2 version 2.5.1 to align and filter Hi-C reads. Unique reads were used as input by CHiCAGO version 1.28.0 (*64*) and significant interactions were called with default parameters.

### GWAS

GWAS were performed using UK Biobank samples. We selected individuals from the White British ancestry set, filtered those with low-quality genotypes (outliers for heterozygosity or missing rate), and excluded individuals with mismatch in their self-reported and genetic sex. We further extracted a subset of unrelated samples by keeping the individual with the smallest number of missing genotypes for every pair of first or second-degree relatives (determined based on genotypes). The following genotype quality controls were applied after sample filtering: minor allele frequency > 0.1%, imputation quality score > 0.8, call rate > 0.95, and Hardy–Weinberg equilibrium exact test p-value > 10^−10^.

We conducted GWAS for the following traits: allergic rhinitis, atopic dermatitis, and COPD. Allergic rhinitis cases were defined as individuals who met any of the following criteria: (1) a report of hay fever, allergic rhinitis, or asthma in field 6152 and a code for allergic rhinitis (1387) in field 20002; (2) an ICD10 code for allergic rhinitis (J30.1, J30.2, J30.3, and J30.4) recorded in fields 41202 or 41204; (3) a doctor-diagnosed allergic rhinitis in field 22126. Atopic dermatitis cases were defined as individuals who met any of the following criteria: (1) a report of hay fever, allergic rhinitis, or asthma in field 6152 and a code for eczema/dermatitis (1452) in field 20002; (2) an ICD10 code for atopic dermatitis (L20.8 and L20.9) in fields 41202 or 41204. COPD cases were defined based on spirometry measurements following the GOLD criteria for moderate to very severe airflow limitation (*65*): individuals with a ratio between FEV1 (field 20150) and FVC (field 20151) less than 0.7 and FEV1 less than 80% of the predicted value (field 20154). For each trait, samples that were not defined as cases were considered as controls.

### Fine-mapping

We followed the pipeline previously described in Zhong, Mitchell, Billstrand, Thompson, Sakabe, Aneas, Salamone, Gu, Sperling, Schoettler, Nobrega, He and Ober (*16*) to perform both standard and functional fine-mapping. Briefly, functional fine-mapping leverages functional annotations to calibrate the prior probabilities of individual SNPs being causal, favoring SNPs located in annotations that contain a higher density of trait-associated variants. Here, we harmonized open chromatin regions (OCRs) from 14 blood and 22 lung cells relevant to AAD pathophysiology, grouped them into five cell lineages (lymphoid, myeloid, epithelial, mesenchymal, and endothelial), and considered OCRs of each lineage as an annotation. For each trait, we ran stratified LD score regression (S-LDSC) to identify lineages that were significantly enriched for trait heritability, conditioning on a set of baseline annotations. OCRs of these lineages were subsequently integrated with GWAS summary statistics for fine-mapping.

To integrate functional annotations with GWAS, we used a hierarchical model, TORUS, to estimate the enrichment of causal variants within each annotation. The relative enrichments between different annotations were then used to estimate a prior causal probability for each SNP (functional prior), based on its overlapped annotations. Finally, we fine-mapped GWAS summary statistics using SuSiE-RSS (*31, 32*), taking functional priors of individual SNPs as input. We assumed 5 independent causal signals at each LD block and set a purity threshold of 0.4. For standard fine-mapping, we applied the default uniform prior while keeping all other parameters unchanged.

### Effector gene scoring

Identification of effector risk genes was performed as previously described (*16*).

### Luciferase assays

Luciferase construct cloning and assays were performed as previously described (*16*) with modifications. Candidate regulatory element sequences were identified as an open chromatin region in ASMC, BEC, IM, or AM containing variant(s) of interest. Sequences were ordered from IDT and cloned into the pGL4.23 plasmid (Promega E8411) using Gibson Assembly Master Mix (NEB E2611). 16HBE cells were transfected using Lipofectamine LTX reagent with PLUS (Life Technologies 15338100). hTERT-immortalized ASMs and THP1s were electroporated using the Lonza 4D-Nucleofector X system with SG cell line kit (Lonza V4XC-3032). To determine haplotype-specific activity between two constructs, Welch’s t test was performed. To determine haplotype-specific activity between three constructs, One-way ANOVA was performed.

## Data availability

All sequencing data is available from Array Express (https://www.ebi.ac.uk/biostudies/arrayexpress) and NCBI’s GEO (https://ncbi.nlm.nih.gov/). Datasets are listed in Supplementary Table 8.

